# Predictors of severe symptomatic laboratory-confirmed SARS-COV-2 reinfection

**DOI:** 10.1101/2020.10.14.20212720

**Authors:** Efrén Murillo-Zamora, Oliver Mendoza-Cano, Iván Delgado-Enciso, Carlos M. Hernandez-Suarez

## Abstract

**Background:** There is a major concern regarding the prognosis of coronavirus disease 2019 (COVID-19) in patients who recovered to first-time illness.

**Objective:** To evaluate factors predicting severe symptomatic laboratory-confirmed (reverse transcription-quantitative polymerase chain reaction, RT-qPCR) SARS-COV-2 (severe acute coronavirus-2) reinfection.

**Method:** We conducted a nationwide retrospective cohort study in Mexico and data from 258 reinfection cases (at least 28 days between both episodes onset) were analyzed. We used risk ratios (RR) and 95% confidence intervals (CI) to evaluate predictors of severe (dyspnea requiring hospital admission) secondary SARS-COV-2 infection.

**Results:** The risk of severe disease was 14.7% and the observed overall fatality rate was 4.3%. Patients with more serious primary disease were more likely to develop severe symptoms (39.5% vs. 5.5%, *p* < 0.001) during reinfection. In multiple analysis, factors associated with an increased risk of severe symptomatic SARS-COV-2 reinfection were increasing age (RR _*per year*_ = 1.007, 95% CI 1.003-1.010), comorbidities (namely obesity [RR = 1.12, 95% CI 1.01-1.24], asthma [RR = 1.26, 95% CI 1.06-1.50], type 2 diabetes mellitus [RR = 1.22, 95% CI 1.07 - 1.38] and previous severe laboratory-confirmed COVID-19 (RR = 1.20, 95% CI 1.03-1.39).

**Conclusions:** To the best of our knowledge this is the first study evaluating disease outcomes in a large set of laboratory-positive cases of symptomatic SARS-COV-2 reinfection and factors associated with illness severity was characterized. Our results may contribute to the current knowledge of SARS-COV-2 pathogenicity and to identify populations at increased risk of a poorer outcome after reinfection.

## Introduction

The COVID-19 (coronavirus disease 2019) by SARS-COV-2 (severe acute respiratory coronavirus 2) pandemic has had a major impact in healthcare systems. In Mexico, by middle October 2020, more than 800 confirmed cases and nearly 85 thousand deaths had been registered [1].

Reinfection risk is a major debated related to the COVID-19 pandemic, even when there is not a consensus regarding its definition [2]. Given the high observed mortality in Mexico, there is concern regarding the impact of subsequent COVID-19 in recovered patients. The aim of this study was to evaluate factors predicting severe SARS-COV-2 symptomatic reinfection in a large and nationwide cohort of laboratory-confirmed COVID-19 survivors.

## Methods

We conducted a nationwide retrospective cohort study including adults (aged 20 years or older) with laboratory-confirmed symptomatic (quantitative reverse-transcription polymerase chain reaction, RT-qPCR; nasopharyngeal or deep nasal swabs were employed) SARS-COV-2 reinfection. The research group previously published a broader description of the employed methods [3].

Subjects with primary COVID-19 onset from March to July 2020, who recovered to severe or non-severe illness (dyspnea requiring hospital admission) were eligible. The main binary outcome was severe symptomatic laboratory-confirmed (RT-qPCR) and it was defined by the reappearance of COVID-19 symptoms at 28 days or above from a primary laboratory-positive COVID-19 [4].

Clinical and epidemiological data of interest were obtained from medical files and death certificates, if applicable. We used linear regression models to calculate risk ratios (RR) and 95% confidence intervals (CI) to evaluate factors associated with the risk of severe symptomatic SARS-COV-2 reinfection. The next models were built: one for each evaluated predictor and a multiple model. The Health Research Committee 601 of the Mexican Institute of Social Security provided approval (R-2020-601-015).

## Results

Data from 100,432 first-time and recovered COVID-19 patients were analyzed and 258 laboratory-confirmed cases of reinfection were identified (0.26%). The study profile is presented as *Supplementary Figure 1*.

Median elapsed days between first and second time COVID-19 were 56 days (interquartile range, IQR, 40-81), and no significant differences were observed between patients with non-severe and severe primary disease (*p* = 0.431). Most of the participants who tested-positive for SARS-COV-2 reinfection were female (53.9%) and were aged 49 years or younger (81.8%). The overall risk of severe subsequent illness was 14.7% (*n* = 38). The observed fatality rate was 4.3% (11/258) and it was higher among patients with severe symptoms (23.3% vs. 4.5%, *p* < 0.001).

When severe and non-severe second-time infections were compared, patients with more serious primary disease were more likely to develop severe symptoms (39.5% vs. 5.5%, *p* < 0.001), as well as those aged 50 years old or above (52.6% vs. 12.3%, *p* < 0.001). In general, the prevalence of chronic non-communicable diseases (namely type 2 diabetes mellitus, arterial hypertension, and kidney disease) was higher among severe subsequent cases. Other characteristics of interest are presented in *Supplementary Table 1*.

In multiple analysis (Table 2), factors associated with a more severe symptomatic SARS-COV-2 reinfection were increasing age (RR _*per year*_ = 1.007, 95% CI 1.003-1.010) and personal history of obesity (RR = 1.12, 95% CI 1.01-1.24), asthma (RR = 1.26, 95% CI 1.06-1.50), type 2 diabetes mellitus (RR = 1.22, 95% CI 1.07 - 1.38) and chronic kidney disease (RR = 1.47, 95% CI 1.21-1.80). Subjects with previous severe COVID-19, had a 20% increased risk (RR = 1.20, 95% CI 1.03-1.39) of also presenting severe symptoms during secondary disease.

## Discussion

The results from our analysis characterized factors determining the risk of severe symptomatic SARS-COV-2 infection. However, and since currently there is not a consensus regarding the definition of SARS-COV-2 reinfection, our findings must be carefully considered. The cut-out points that we used, that corresponds to one of the criteria proposed by *Tomassini et al*. [2], seems highly plausible since it is based in the observed viral decay in laboratory-positive cases of COVID-19 [5]. All the enrolled subjects from our study became asymptomatic between both episodes.

We would like to highlight that enrolled patients who recovered to severe primary illness had a 20% increase in the risk of severe subsequent disease (RR = 1.20, 95% CI 1.03-1.39). Besides, a shorter interval between both disease episodes was documented in patients with severe primary illness (median days [IQR]: 48 [33-62] vs. 57 [41-84]; *p* = 0.040).

If later replicated in other populations, this finding may be highly relevant for public health policymakers since it would imply that: i) even when higher antibodies titers have been documented among patients with severe COVID-19 [5], these would not protect them from a secondary severe illness about 4-8 weeks later and ii) these initially recovered patients would benefit from particularly strict measures to prevent the spread of respiratory viral pathogens. However, in our study sample, the risk of second-time disease seemed to be low (0.26%).

The rest of the factors predicting severe symptomatic SARS-COV-2 reinfection (namely increasing age, obesity, asthma, type 2 diabetes mellitus, and chronic kidney disease) did not seem to differ from those determining a severe primary disease [6]. These findings support that medical and control interventions in subsequent COVID-19 cases, in general terms, do not seem to differ from primary infections [7].

Interestingly, most of the laboratory-positive second-time cases were young (under 50 years old). A reduced COVID-19 awareness among younger subjects may be implied [8], particularly due to a high prevalence (87.0%) of non-severe primary illness in this age group.

The potential limitations of this study must be cited. First, the utilized definition of severe COVID-19 in our study represents a limitation that must be cited given that no laboratory-related data (e.g. PaO_2_) was collected. However, a meta-analysis including 15 studies evidenced that dyspnea is a stand-alone prognostic factor of COVID-19-related mortality [9]. Besides, and according to normative standards that have been followed since the beginning of the pandemic in Mexico, hospitalization criteria include the use of validated scales as PSI (Pneumonia Severity Index) and CURB-65 (Confusion, Urea level, Respiratory rate, Blood pressure, and age ≥ 65) [10]. Therefore, this severity indicator seems to be plausible in the study population.

Second, since no mass screening has been performed in Mexico, we were unable to identify second-time asymptomatic cases of SARS-COV-2 infection. Moreover, an undetermined fraction of patients that are currently being identified as primary COVID-19 may correspond to subsequent infections and the related public health implications are unknown.

## Conclusions

We identified factors predicting severe symptomatic SARS-COV-2 reinfection, which seemed to be a rare event. To the best of our knowledge, this is the first study aiming to characterize clinical outcomes in patients with second time COVID-19. If later replicated, our results may be highly useful for implementing interventions focusing on the reduction of disease burden in populations at risk, including vaccination efforts.

## Data Availability

Data may be available after the request to corresponding author.

## Tables and Figures

**Table 1.** Predictors of severe symptomatic laboratory-confirmed SARS-COV-2 reinfection, Mexico 2020

|  | RR (95% CI), *p* |  |  |  |  |  |
| --- | --- | --- | --- | --- | --- | --- |
|  | Bivariate analysis |  |  | Multiple analysis |  |  |
| **Gender** |  |  |  |  |  |  |
| Female | 1.00 |  |  | 1.00 |  |  |
| Male | 1.06 | (0.97 - 1.15) | 0.221 | 1.02 | (0.95 - 1.10) | 0.589 |
| **Age group (years)** |  |  |  |  |  |  |
| 20-49 | 1.00 |  |  | 1.00 |  |  |
| 50-59 | 1.36 | (1.21 - 1.54) | < 0.001 | 1.22 | (1.09 - 1.37) | 0.001 |
| 60-69 | 1.41 | (1.10 - 1.81) | 0.007 | 1.18 | (0.93 - 1.49) | 0.176 |
| 70+ | 1.63 | (1.27 - 2.09) | < 0.001 | 1.17 | (0.89 - 1.51) | 0.262 |
| **Disease severity**<br>**(at primary infection) <sup>a</sup>** |  |  |  |  |  |  |
| Mild - moderate | 1.00 |  |  | 1.00 |  |  |
| Severe | 1.58 | (1.38 - 1.80) | < 0.001 | 1.20 | (1.03 - 1.39) | 0.019 |
| *Personal history of:* |  |  |  |  |  |  |
| **Obesity (BMI 30 or higher)** |  |  |  |  |  |  |
| No | 1.00 |  |  | 1.00 |  |  |
| Yes | 1.10 | (0.97 - 1.24) | 0.131 | 1.12 | (1.01 - 1.24) | 0.025 |
| **Asthma** |  |  |  |  |  |  |
| No | 1.00 |  |  | 1.00 |  |  |
| Yes | 1.22 | (0.99 - 1.49) | 0.062 | 1.26 | (1.06 - 1.50) | 0.009 |
| **Type 2 diabetes mellitus** |  |  |  |  |  |  |
| No | 1.00 |  |  | 1.00 |  |  |
| Yes | 1.52 | (1.34 - 1.72) | < 0.001 | 1.22 | (1.07 - 1.38) | 0.003 |
| **Immunosuppression <sup>b</sup>** |  |  |  |  |  |  |
| No | 1.00 |  |  | 1.00 |  |  |
| Yes | 1.02 | (0.76 - 1.36) | 0.893 | 0.91 | (0.72 - 1.16) | 0.467 |
| **Chronic kidney disease** |  |  |  |  |  |  |
| No | 1.00 |  |  | 1.00 |  |  |
| Yes | 1.88 | (1.55 - 2.28) | < 0.001 | 1.47 | (1.21 - 1.80) | < 0.001 |

Abbreviations: **RR**, Risk ratio; **CI**, Confidence interval; **BMI**, Body mass index.
Notes: 1) Generalized linear regression models were used to obtain RR and 95% CI; 2) Multiple regression coefficients were adjusted by variables listed in the table.
<sup>a</sup> Severe illness included the register of dyspnea requiring hospital admission.
<sup>b</sup> Immunosuppression referred to any cause of the related deficiency except for type 2 diabetes mellitus or renal impairment.

**Supplementary Figure 1.**
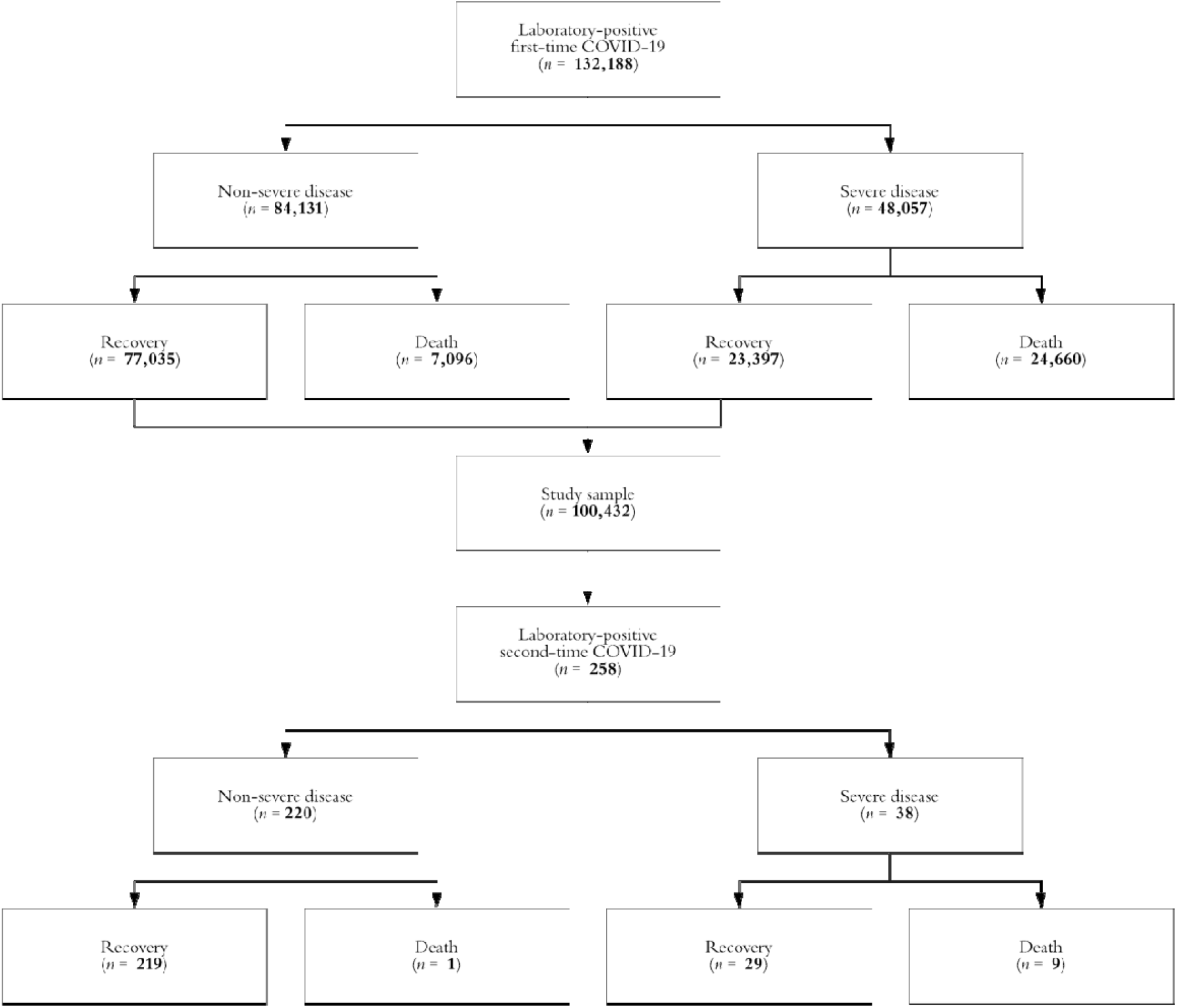
Study profile, Mexico 2020 Abbreviations: **COVID-19**, Coronavirus disease 2019. Note: The definition of severe first or second-time disease included the presence of dyspnea requiring hospital admission.

**Supplementary Table 1.**
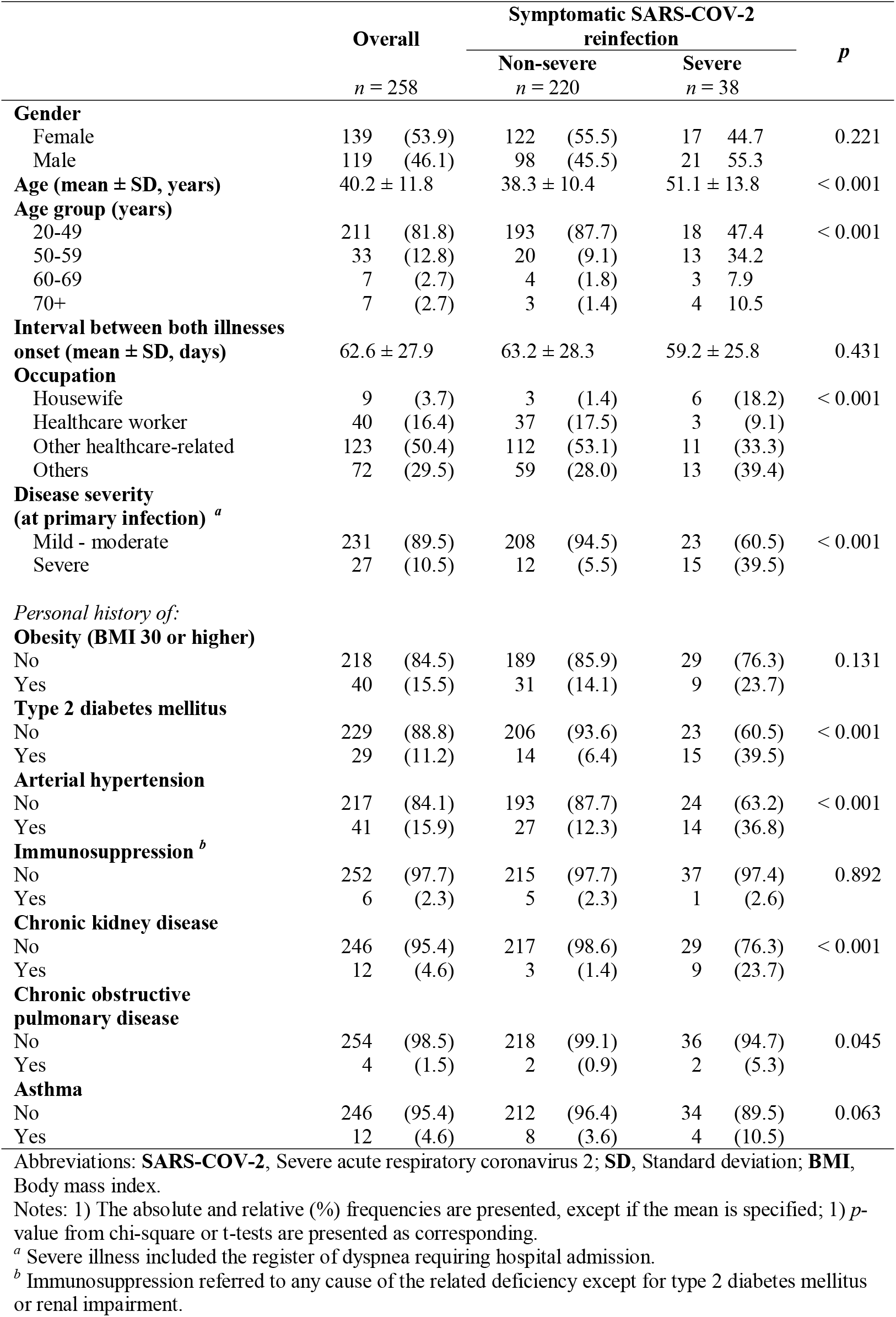
Characteristics of the study sample according to disease severity, Mexico 2020

|  | Overall |  | Symptomatic SARS-COV-2 reinfection |  |  |  | p |
| --- | --- | --- | --- | --- | --- | --- | --- |
|  | n = 258 |  | Non-severe<br>n = 220 |  | Severe<br>n = 38 |  |  |
| **Gender** |  |  |  |  |  |  |  |
| Female | 139 | (53.9) | 122 | (55.5) | 17 | 44.7 | 0.221 |
| Male | 119 | (46.1) | 98 | (45.5) | 21 | 55.3 |  |
| **Age (mean ± SD, years)** | 40.2 ± 11.8 |  | 38.3 ± 10.4 |  | 51.1 ± 13.8 |  | < 0.001 |
| **Age group (years)** |  |  |  |  |  |  |  |
| 20-49 | 211 | (81.8) | 193 | (87.7) | 18 | 47.4 | < 0.001 |
| 50-59 | 33 | (12.8) | 20 | (9.1) | 13 | 34.2 |  |
| 60-69 | 7 | (2.7) | 4 | (1.8) | 3 | 7.9 |  |
| 70+ | 7 | (2.7) | 3 | (1.4) | 4 | 10.5 |  |
| **Interval between both illnesses onset (mean ± SD, days)** | 62.6 ± 27.9 |  | 63.2 ± 28.3 |  | 59.2 ± 25.8 |  | 0.431 |
| **Occupation** |  |  |  |  |  |  |  |
| Housewife | 9 | (3.7) | 3 | (1.4) | 6 | (18.2) | < 0.001 |
| Healthcare worker | 40 | (16.4) | 37 | (17.5) | 3 | (9.1) |  |
| Other healthcare-related | 123 | (50.4) | 112 | (53.1) | 11 | (33.3) |  |
| Others | 72 | (29.5) | 59 | (28.0) | 13 | (39.4) |  |
| **Disease severity (at primary infection) <sup>a</sup>** |  |  |  |  |  |  |  |
| Mild - moderate | 231 | (89.5) | 208 | (94.5) | 23 | (60.5) | < 0.001 |
| Severe | 27 | (10.5) | 12 | (5.5) | 15 | (39.5) |  |
| *Personal history of:* |  |  |  |  |  |  |  |
| **Obesity (BMI 30 or higher)** |  |  |  |  |  |  |  |
| No | 218 | (84.5) | 189 | (85.9) | 29 | (76.3) | 0.131 |
| Yes | 40 | (15.5) | 31 | (14.1) | 9 | (23.7) |  |
| **Type 2 diabetes mellitus** |  |  |  |  |  |  |  |
| No | 229 | (88.8) | 206 | (93.6) | 23 | (60.5) | < 0.001 |
| Yes | 29 | (11.2) | 14 | (6.4) | 15 | (39.5) |  |
| **Arterial hypertension** |  |  |  |  |  |  |  |
| No | 217 | (84.1) | 193 | (87.7) | 24 | (63.2) | < 0.001 |
| Yes | 41 | (15.9) | 27 | (12.3) | 14 | (36.8) |  |
| **Immunosuppression <sup>b</sup>** |  |  |  |  |  |  |  |
| No | 252 | (97.7) | 215 | (97.7) | 37 | (97.4) | 0.892 |
| Yes | 6 | (2.3) | 5 | (2.3) | 1 | (2.6) |  |
| **Chronic kidney disease** |  |  |  |  |  |  |  |
| No | 246 | (95.4) | 217 | (98.6) | 29 | (76.3) | < 0.001 |
| Yes | 12 | (4.6) | 3 | (1.4) | 9 | (23.7) |  |
| **Chronic obstructive pulmonary disease** |  |  |  |  |  |  |  |
| No | 254 | (98.5) | 218 | (99.1) | 36 | (94.7) | 0.045 |
| Yes | 4 | (1.5) | 2 | (0.9) | 2 | (5.3) |  |
| **Asthma** |  |  |  |  |  |  |  |
| No | 246 | (95.4) | 212 | (96.4) | 34 | (89.5) | 0.063 |
| Yes | 12 | (4.6) | 8 | (3.6) | 4 | (10.5) |  |

Abbreviations: **SARS-COV-2**, Severe acute respiratory coronavirus 2; **SD**, Standard deviation; **BMI**, Body mass index.
Notes: 1) The absolute and relative (%) frequencies are presented, except if the mean is specified; 1) *p*-value from chi-square or t-tests are presented as corresponding.
<sup>a</sup> Severe illness included the register of dyspnea requiring hospital admission.
<sup>b</sup> Immunosuppression referred to any cause of the related deficiency except for type 2 diabetes mellitus or renal impairment.

